# Comparison of common outcome measures for assessing independence in patients diagnosed with disorders of consciousness: A Traumatic Brain Injury Model Systems Study

**DOI:** 10.1101/2022.02.04.22270464

**Authors:** Samuel B. Snider, Robert G. Kowalski, Flora Hammond, Saef Izzy, Shirley L. Shih, Craig Rovito, Brian L. Edlow, Ross D. Zafonte, Joseph T. Giacino, Yelena G. Bodien

## Abstract

**Importance:** Patients with disorders of consciousness (DoC) after traumatic brain injury (TBI) recover to varying degrees of functional dependency. Dependency is difficult to measure but critical for outcome interpretation and prognostic counseling. Traditional outcome measures, like the Glasgow Outcome Scale-Extended (GOSE), are mandated by the US Food and Drug Administration for evaluating TBI clinical trial efficacy but have an unknown accuracy for measuring dependency.

**Objective:** We used the Functional Independence Measure (FIM®) as the reference standard to evaluate how accurately the GOSE and Disability Rating Scale (DRS) assess functional dependency in the world’s largest cohort of patients with DoC after TBI. We propose an alternate, data-driven, approach to measuring dependency.

**Design, Setting, and Participants:** In this cohort study, we included patients with DoC prospectively enrolled in the longitudinal Traumatic Brain Injury Model Systems National Database (TBIMS NDB). Participants were survivors of moderate/severe TBI with DoC on admission to a US inpatient rehabilitation center between 1988 and 2020, followed 1 year after injury.

**Exposure:** We examined the classification performance of common TBI outcome measure cutpoints (GOSE ≤3 and ≤4 [Lower and Upper Severe Disability, respectively], and DRS ≥12 [Severe Disability]) in identifying subjects with functional dependency at 1 year. We compared data-derived optimal cut-points on these scales to a novel DRS-based marker of dependency, the DRS_Depend_.

**Main Outcome and Measure:** Total FIM score < 80 (FIM-dependency) at 1 year.

**Results:** Of 18,486 TBIMS participants, 1,483 with DoC on arrival to inpatient rehabilitation met inclusion criteria (mean [SD] age=38 [18] years; 76% male). The sensitivity of GOSE cut-points of ≤3 and ≤4 for identifying FIM-dependency were 97% and 98%, but specificities were 73% and 51%, respectively. The sensitivity of the DRS cut-point of ≥12 was 60%, but specificity was 100%. The DRS_Depend_ had a sensitivity of 83% and a specificity of 94% for classifying FIM-dependency, with a greater AUROC than the data-derived optimal GOSE (≤3, p=0.01) and DRS (≥10, p=0.008) cut-points.

**Conclusions and Relevance:** Commonly-used GOSE and DRS cut-points have limited sensitivity or specificity for identifying functional dependency. The DRS_Depend_ identifies FIM-dependency more accurately than GOSE and DRS cut-points, but requires further validation.

## INTRODUCTION

Most patients with a disorder of consciousness (DoC) after traumatic brain injury (TBI) ultimately recover consciousness^1-3^. However, studies of short^1^, medium^2^, and long-term^3-5^ outcomes in this patient population suggest that levels of function vary widely, from independent and employable to dependent for all basic needs. TBI clinical trials track recovery using standardized outcome measures, such as the Glasgow Outcome Scale - Extended (GOSE) ^6, 7^ or Disability Rating Scale (DRS)^8^, and often collapse the distribution of scores into ‘favorable’ and ‘unfavorable’ groups to simplify analysis and interpretation. The precise cut-points used to generate these groups differ across measures and across trials. While there is no universal definition for ‘unfavorable’ outcome, in a recent study, most families of patients with severe brain injury in the intensive care unit reported that complete dependency, defined as requiring assistance with all physical and cognitive tasks, would not be an acceptable long-term outcome^9^. The accuracy of different GOSE and DRS cut-points for identifying complete dependency in patients recovering from TBI is unknown.

The Functional Independence Measure (FIM®)^10^, a comprehensive instrument that scores level of assistance needed across 13 physical and 5 cognitive dimensions, provides a granular assessment of the severity of a patient’s functional impairment. The FIM is used extensively as a research outcome measure in the rehabilitation setting and has been studied across the trajectory of recovery from non-neurologic and neurologic injury and illness, including TBI^11, 12^. FIM scores reflect the time (hours per day) and type (supervision versus assistance) of external support a patient requires^13, 14^ to complete basic tasks. A FIM total score less than 80 indicates complete dependence (i.e., FIM-dependency) and reflects a burden of care that cannot typically be provided in the home^15-18^. The FIM is not commonly used in interventional TBI clinical trials, most established TBI outcome databases, or other medical disciplines, and is being phased out of routine use.

The Glasgow Outcome Scale - Extended (GOSE)^6, 7^, an 8-item ordinal scale, is the most frequently used TBI outcome assessment and the only measure accepted by the US Food and Drug Administration as a primary outcome in TBI trials^19^. However, each GOSE level encompasses a wide range of function and clinically meaningful differences in disability may be missed^20, 21^. An alternative to the GOSE, the Disability Rating Scale (DRS)^8^, was designed to reflect the full range of outcomes after TBI, from coma to return to competitive employment^19, 22^. Limitations of the DRS include lack of precision and ceiling effects^4, 20^. Though both the GOSE and the DRS have individual items evaluating dependence, how well these measures align with the FIM, which was designed and validated to directly measure dependence, has not been established.

Using data from the Traumatic Brain Injury Model Systems (TBIMS) National Database (NDB), the largest prospective, longitudinal TBI cohort in the world^23^, we compared multiple methods of characterizing 1-year functional dependence in participants admitted to inpatient rehabilitation with DoC. Our primary aims were to: 1) characterize and compare the score distribution of three disability scales: FIM, GOSE, and DRS and 2) assess the accuracy of GOSE and DRS dichotomization cut-points for identifying participants scoring in the dependent range on the FIM (i.e., FIM-dependency, total score <80). In exploratory analyses, we used a data-driven approach to derive optimal GOSE and DRS cut-points for classifying FIM-dependency. Finally, we developed a simple, binary dependency score derived from a subset of DRS items, (the DRS_Depend_ Score), and compared this measure with the FIM and the GOSE-E and DRS total-score cut-points.

## METHODS

We conducted a retrospective analysis of a prospective longitudinal cohort study of participants enrolled in the TBIMS NDB^23^. Characteristics of this continuously enrolling database have been described previously^1, 24, 25^. The sample includes participants who survived acute hospitalization and were admitted to one of 16 inpatient rehabilitation centers in the US that participated in the TBIMS program. This study was approved by the Massachusetts General Brigham institutional review board (IRB Protocol #2012P002476) and by IRBs at each participating center in the TBIMS NDB.

### TBIMS Procedures and Study Sample

As described previously, TBIMS participants are survivors of acute moderate or severe TBI and 16 years or older at injury onset^1, 26^. For this study, we included participants admitted to an inpatient rehabilitation facility with DoC, defined here and in prior studies^1, 3^ as an admission DRS motor item score >0 (i.e., absence of command following). We excluded participants who did not have a FIM score and either the DRS or GOSE acquired at 1-year (365 +/-60 days post-injury, as recommended by the TBIMS standard operating procedure)^26^. Among 18,486 participants in the TBIMS database enrolled between 10/1/1988 and 09/04/2020, our final cohort included 1,483 participants with DoC who met inclusion criteria (Figure 1).

**Figure 1:**
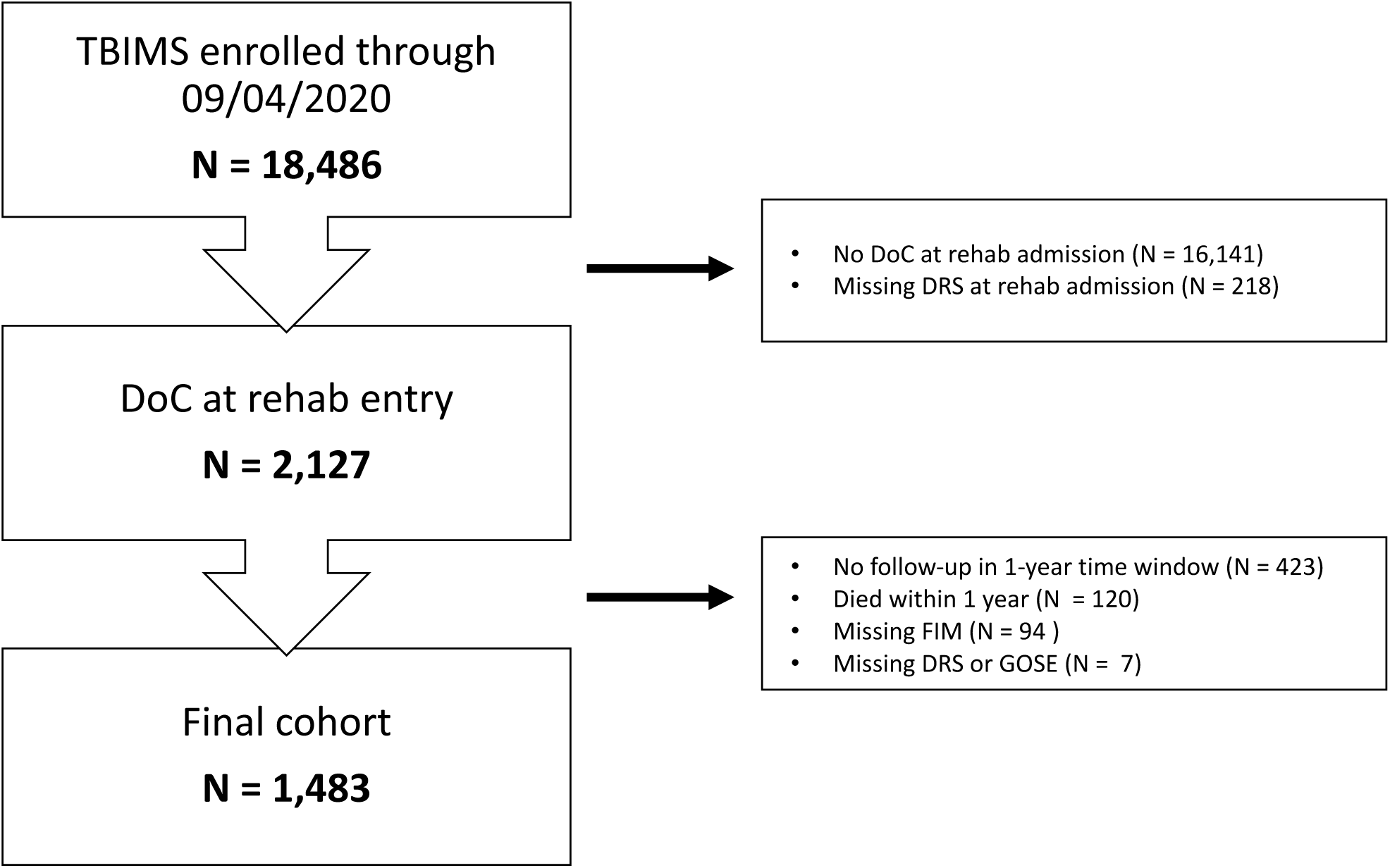
Cohort description. Study CONSORT diagram. Abbreviations: TBIMS = Traumatic Brain Injury Model Systems. DoC = Disorders of Consciousness, DRS = Disability Rating Scale, FIM = Functional Independence Measure, GOSE = Glasgow Outcome Scale Extended.

### Measures

The TBIMS NDB includes demographics and TBI characteristics, as well as acute care hospital, rehabilitation, and follow-up outcome variables^1, 26^. Follow-up assessments are conducted at 1 year, 2 years, 5 years, and every 5 years after injury until death or withdrawal from the study. Study investigators are trained to collect data from the medical record and to perform all assessments in accordance with quality control guidelines maintained by the TBIMS National Data and Statistical Center^26^. We selected the 1-year time-point for analysis, as it was previously shown to reflect the range of longer-term outcomes in TBIMS participants with DoC on rehabilitation admission^3^.

#### FIM

The FIM instrument includes a series of standardized questions assessing the degree of caregiver support required to accomplish basic cognitive and motor tasks, with total scores ranging from 18 (most dependent) to 126 (least dependent)^16^. Based on external validation studies^13-15, 18^ and recent observational studies in this cohort^1^, we defined functional dependency as FIM total score <80 (FIM-dependency). We used the FIM as our study reference standard because of its granularity and extensive validation in multiple previous investigations ^13-15, 18^.

#### GOSE

The GOSE is an 8-point scale ranging from 1 (Death) to 8 (Upper Good Recovery), based on degree of functional difficulties in major life domains^20, 27, 28^. Though the GOSE can be analyzed ordinally^29^, it is often dichotomized into ‘favorable’ and ‘unfavorable’ outcomes, with an ‘unfavorable’ cut-point set at ≤3 (Lower Severe Disability, e.g. *cannot* be left unsupervised in the home for more than 8 hours)^2, 30-32^ or ≤4 (Upper Severe Disability, e.g. *can* be left unsupervised in the home for more than 8 hours, but dependent outside the home)^33-35^. We assessed the performance of these cut-points and determined a data-derived optimal threshold for classifying FIM-dependency.

#### DRS

The DRS combines the three items of the Glasgow Coma Scale^36^ with five additional measures^8^. These additional measures evaluate a subject’s awareness of how and when to feed, groom, and toilet (0: complete awareness to 3: no awareness), level of functional dependence on others (0: completely independent to 5: totally dependent), and employability (0: not restricted to 3: not employable). The eight item scores are then summed to generate a total score of 0 (least disabled) to 30 (dead)^8^. Prior observational and interventional studies^2, 37^ have categorized DRS total scores into ad-hoc groupings such as: None (0), Mild (1), Partial (2-3), Moderate (4-6), Moderately-severe (7-11), Severe (12-16), Extremely severe (17-21), Vegetative state (22 -24), Extreme vegetative state (25-29) ^8^. We evaluated the performance of the Severe Disability cutpoint (scores ≥ 12), as well as a data-driven optimal cut-point for classifying FIM-dependency.

#### DRS_*Depend*_

Based on the standardized survey instrument, the GOSE cut-points of ≤3 and ≤4 represent some degree of in-home dependence^38^, though not necessarily resulting from cognitive impairment. The DRS cut-point of Severe Disability (score ≥ 12) cannot be directly mapped onto level of dependence. We generated a binary DRS rating (the DRS_Depend_) using a subset of DRS items and tested whether this rating classified FIM-dependency better than the GOSE and total DRS. We defined DRS_*Depend*_ as a DRS profile with Level of Functioning ≥ 4 (indicating the need for assistance with all activities at all times), *and* a score of > 0 (some assistance needed) on at least one of the following items: Verbal, Feeding, Toileting, or Grooming. Participants meeting *DRS_Depend_* criteria, therefore, require the assistance of another person at all times, and that need is based, at least partially, on cognitive impairment. We evaluated the performance of the DRS_*Depend*_ for classifying FIM-dependency.

### Statistical Analysis

First, we compared baseline demographics and injury characteristics of participants who completed the 1-year follow-up assessment (N=1,483) to those who missed the 1-year follow-up or died (N=644) using T (continuous, normally distributed variables), Wilcoxon (continuous, nonnormally distributed variables) or *χ*^2^ (categorical variables) tests (Table 1).

**Table 1:** Demographic and Injury Characteristics of Participants Diagnosed with DoC on Admission to Rehabilitation

|  |  |  |  |
| --- | --- | --- | --- |
| **Variable** | **1-yr Outcome Complete**<br>**(N = 1,483)** | **Deceased or 1-year Outcome Missing**<br>**(N = 644)** | **P value** |
| **Age (Mean [SD])** | 38 [18] | 43 [20] | **< 0.001** |
| Sex (% Male) | 76 | 76 | 0.9 |
| Race (% White) | 68 | 67 | 0.5 |
| **Injury Mechanism (% high velocity)** | **59** | **45** | **< 0.001** |
| Intubated on arrival (%) | 55 | 52 | 0.2 |
| Initial GCS (Mean [SD])<sup>a</sup> | 8 [4] | 9 [4] | 0.1 |
| Craniectomy performed (%) | 20 | 20 | 0.9 |
| **Any Intracranial hypertension (%)** | **43** | **37** | **0.007** |
| SDH or SAH (%) | 99 | 99 | 1 |
| Any contusion (%) | 79 | 81 | 0.5 |
| **IVH (%)** | **44** | **38** | **0.01** |
| Spinal cord injury (%) | 4 | 4 | 0.8 |
| GCS on arrival to rehab (Mean [SD]) | 10 [2] | 10 [3] | 0.2 |
| DRS on arrival to rehab (Median [IQR]) | 22 [19, 23] | 21 [19, 24] | 0.6 |
| **DRS on rehab discharge (Median [IQR])** | **9 [6, 14]** | **10 [6, 16]** | **0.003** |
| Acute Care Length of Stay (Mean Days [SD]) | 30 [21] | 29 [19] | 0.5 |
| Time from injury to 1-yr outcome assessment (Mean days [SD]) | 370 [36] | -- | -- |

a: 57% un-scorable (due to intubation + sedation/paralytic). Bold text indicated a significant difference between groups
**Abbreviations:** TBI traumatic brain injury, DoC Disorders of Consciousness, SD standard deviation, GCS Glasgow Coma Scale, SDH subdural hemorrhage, IVH intraventricular hemorrhage, SAH subarachnoid hemorrhage, DRS Disability Rating Scale

For the primary analysis, we measured performance characteristics for DRS and GOSE cut-points for classifying participants meeting our reference standard criteria for FIM-dependency. For each cut-point, we computed classification performance measures, including area under the receiver operating characteristic curve (AUROC), sensitivity, specificity, positive predictive value (PPV), and negative predictive value (NPV), using 95% confidence intervals generated from 1000 bootstrapped samples, with performance assessed in the out-of-bag sample (R package: cutpointr)^39^. An applied definition of each performance measure is provided in Supplementary Table 1.

In an exploratory analysis, we defined the optimal GOSE and DRS-based thresholds for identifying FIM-dependency. We serially assessed every possible GOSE and DRS dichotomization cut-point, finding the value for each scale that minimized the number of FIM misclassifications. We based our analysis on the assumption that false positives (inappropriately classifying an independent patient as dependent) represent an error with more clinical consequences than false negatives (inappropriately classifying a dependent patient as independent), and assigned a 2:1 false positive-to-false negative misclassification penalty (R: cutpointr)^39^. We then compared the resulting AUROC of these data-driven thresholds to the DRS_Depend_ using Delong’s Tests.

## RESULTS

### Study Population

Within the TBIMS database (N=18,486), 2,127 participants met criteria for DoC on arrival to inpatient rehabilitation; 1,483 survived and were assessed with the FIM and either a DRS or GOSE at 1-year post-injury (Figure 1). Characteristics of the participants meeting criteria for DoC at rehabilitation admission are listed in Table 1. Compared to those who died or had missing outcome data at 1-year (N=644), participants with follow-up (N=1,483) were younger (95% CI for age difference: -3 to -6 years, p<0.001), had less severe DRS scores at rehabilitation discharge (95% CI for score difference: -2 to 0, p<0.001), and were more likely to have had a high velocity injury mechanism, intracranial hypertension, and intraventricular hemorrhage (Table 1).

### Score Distributions for the FIM, GOSE, and DRS

The FIM, GOSE, and DRS total scores were non-normally distributed (all p for Shapiro-Wilk’s tests <0.001). At 1-year post-injury, different proportions of participants met criteria for FIM-dependency (total score < 80, 24%, Figure 2A), Lower Severe Disability on the GOSE (scores ≤3, 44%, Figure 2B), and Severe Disability on the DRS (scores ≥12, 15%, Figure 2; *χ*^2^ =312, p<0.001). While the FIM (median [1^st^ quartile, 3^rd^ quartile]: 112 [83, 121], mode: 126, *Independent*) and DRS (median: 5 [2, 8], mode: 0, *No Disability*) distributions skewed toward the no disability scale extremes, the GOSE distribution skewed toward the severe disability scale extreme (median 4: [3, 6], mode: 3, *Lower Severe Disability*).

**Figure 2:**
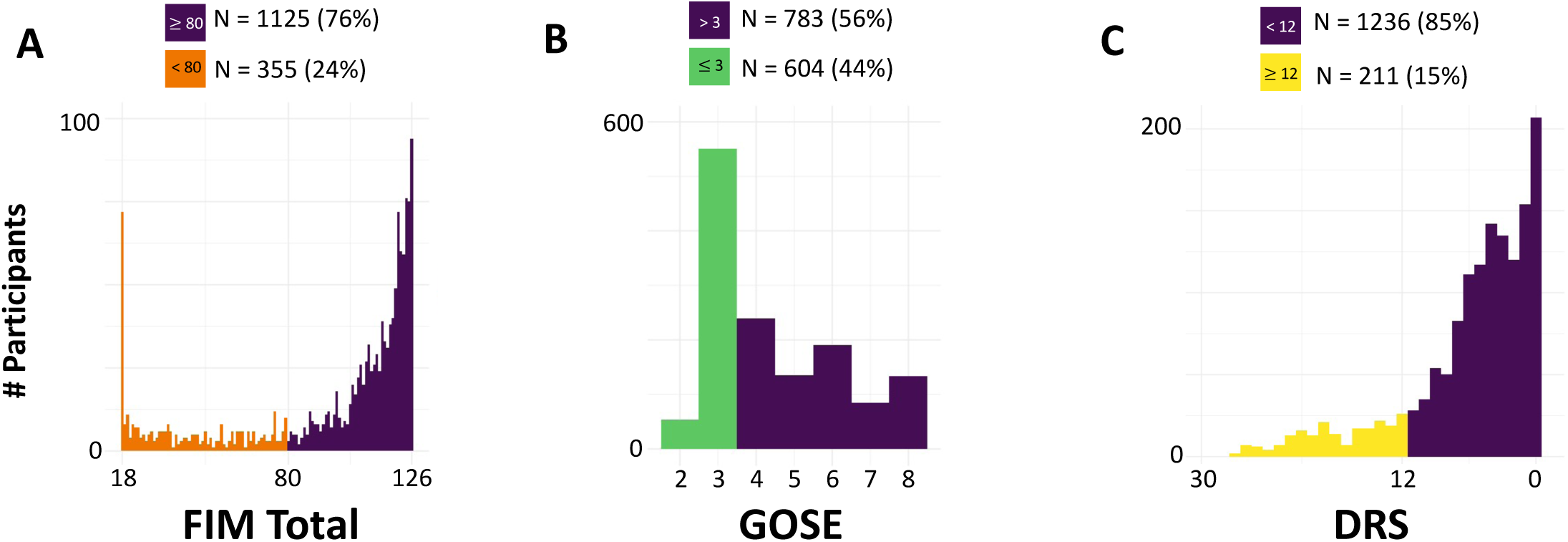
1-year Outcome Distribution Differs by Disability Scale. Histogram of number of patients with each score on the (A) Functional Independence Measure (FIM), (B) Glasgow Outcome Scale Extended (GOSE), (C) Disability Rating Scale (DRS) at 1 year post injury. Orange color (A) indicates FIM-dependency (FIM total score < 80), Green (B) indicates GOSE score ≤ 3 (Lower Severe Disability or worse), and yellow (C) indicates DRS ≥ 12 (Severe Disability or worse).

### Cross-scale correlations and performance characteristics

We next evaluated cross-scale correlations and the performance of different GOSE and DRS thresholds in classifying participants with FIM-dependency.

#### GOSE and FIM

Although there was a significant correlation between GOSE and FIM total scores (Spearman Rho =0.8, p<0.001), total FIM scores of participants with the most common GOSE score (i.e., 3, Lower Severe Disability) were distributed across the full range of the FIM (Figure 3A). The performance characteristics of the GOSE ≤3 and GOSE ≤4 cut-points for classifying FIM-dependency are shown in Table 2. While both cut-points had sensitivities and NPVs for FIM-dependency of at least 97%, the specificities were 74% (bootstrapped 95% CI: [71% - 77%], GOSE ≤3) and 51% ([48% - 54%], GOSE ≤4), and PPVs were 54% ([50% - 59%], GOSE ≤3) and 39% ([36% - 43%], GOSE ≤4). Though 99% of participants with GOSE scores greater than 3 or 4 *did not* meet FIM- dependency criteria, only 54% of participants with GOSE scores ≤3, and 39% of participants with GOSE scores ≤4, *met* FIM-dependency criteria.

**Figure 3:**
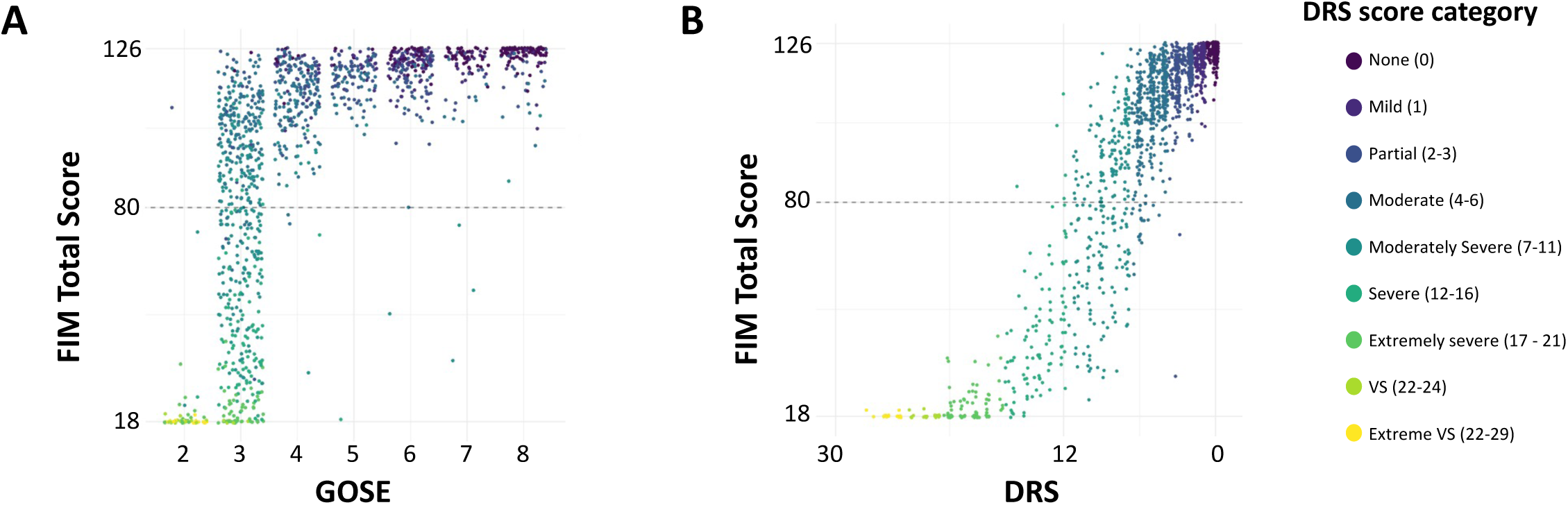
Distribution of GOSE and DRS Scores Compared to the FIM at 1-year Post Injury. Glasgow Outcome Scale-Extended (GOSE) scores (A) and Disability Rating Scale (DRS) scores (B) are plotted against Functional Independence Measure (FIM) total scores. In both panels, points are randomly jittered to avoid overlap and colored by their DRS ordinal category. The dashed line signifies a FIM total score of 80.

**TABLE 2:** Performance of GOSE and DRS Cut-Points for Identifying FIM-dependent Participants

|  |  |  |  |  |  |
| --- | --- | --- | --- | --- | --- |
| **Cut-point** | **AUROC mean**<br>**[95% CI]** | **Sensitivity mean**<br>**[95% CI]** | **Specificity mean**<br>**[95% CI]** | **PPV mean**<br>**[95% CI]** | **NPV mean**<br>**[95% CI]** |
| GOSE $\leq 3^{a,b}$ | 0.85<br>[0.84 – 0.87] | 97%<br>[95 – 99] | 74%<br>[71 – 77] | 54%<br>[50 – 59] | 99%<br>[98 – 100] |
| GOSE $\leq 4^a$ | 0.75<br>[0.73 – 0.77] | 98%<br>[97 – 100] | 51%<br>[48 – 54] | 39%<br>[36 – 43] | 99%<br>[98 – 100] |
| DRS $\geq 12^a$ | 0.80<br>[0.77 – 0.83] | 60%<br>[54 – 66] | 100%<br>[99 – 100] | 98%<br>[95 – 100] | 89%<br>[87 – 91] |
| DRS $\geq 10^b$ | 0.86<br>[0.83 – 0.88] | 74%<br>[69 – 79] | 98%<br>[97 – 99] | 92%<br>[88 – 95] | 92%<br>[91 – 94] |
| DRS<sub>Depend</sub> | 0.89<br>[0.86 – 0.91] | 83%<br>[78 – 87] | 94%<br>[93 – 96] | 82%<br>[78 – 87] | 95%<br>[93 – 96] |

a = literature-derived cut-point
b = data-derived cut-point
Abbreviations: PPV positive predictive value, NPV negative predictive value, GOSE Glasgow Outcome Scale Extended, DRS Disability Rating Scale

#### DRS and FIM

The DRS and the FIM were also correlated (Spearman Rho =-0.9, p<0.001, Figure 3B). Similar to participants with GOSE scores of 3, we found that subjects with modal DRS scores (Moderate Disability, total scores: 4-6) spanned a wide range of FIM scores (Figure 3B, blue-green). While the DRS cut-point of Severe Disability or worse (DRS ≥12) had specificity and PPV for FIM-dependency of greater than 98%, the sensitivity was 60% [54% - 66%] and the NPV was 89% [87% - 91%]). Though 98% of participants with DRS scores ≥12 met criteria for FIM-dependency, only 60% of participants who met criteria for FIM-dependency also had DRS scores ≥12.

### Exploratory Analyses

The data-derived optimal cut-points for identifying FIM-dependency were GOSE scores ≤3 and total DRS scores ≥10 (Table 2). As measured by the AUROC, there was no difference in the discriminative capacity between the optimal GOSE (GOSE ≤3: AUROC 0.85 [0.84 – 0.87]) and DRS (DRS ≥10: AUROC 0.86 [0.83 – 0.88]) cut-points (p=0.7)

### DRS_Depend_

We assessed the classification performance of a DRS-based functional dependency metric, the DRS_Depend_. The DRS_Depend_ classified FIM-dependency with 83% [78% - 87%] sensitivity, 94% [93% - 96%] specificity, PPV of 82% [78% - 87%], NPV of 95% [93% - 96%] and an AUROC of 0.89 [0.86 – 0.91] (Table 2, Figure 4). The DRS_Depend_ discriminated FIM-dependency better than the data-derived optimal GOSE (p=0.01) and DRS (p=0.008) cut-points.

**Figure 4:**
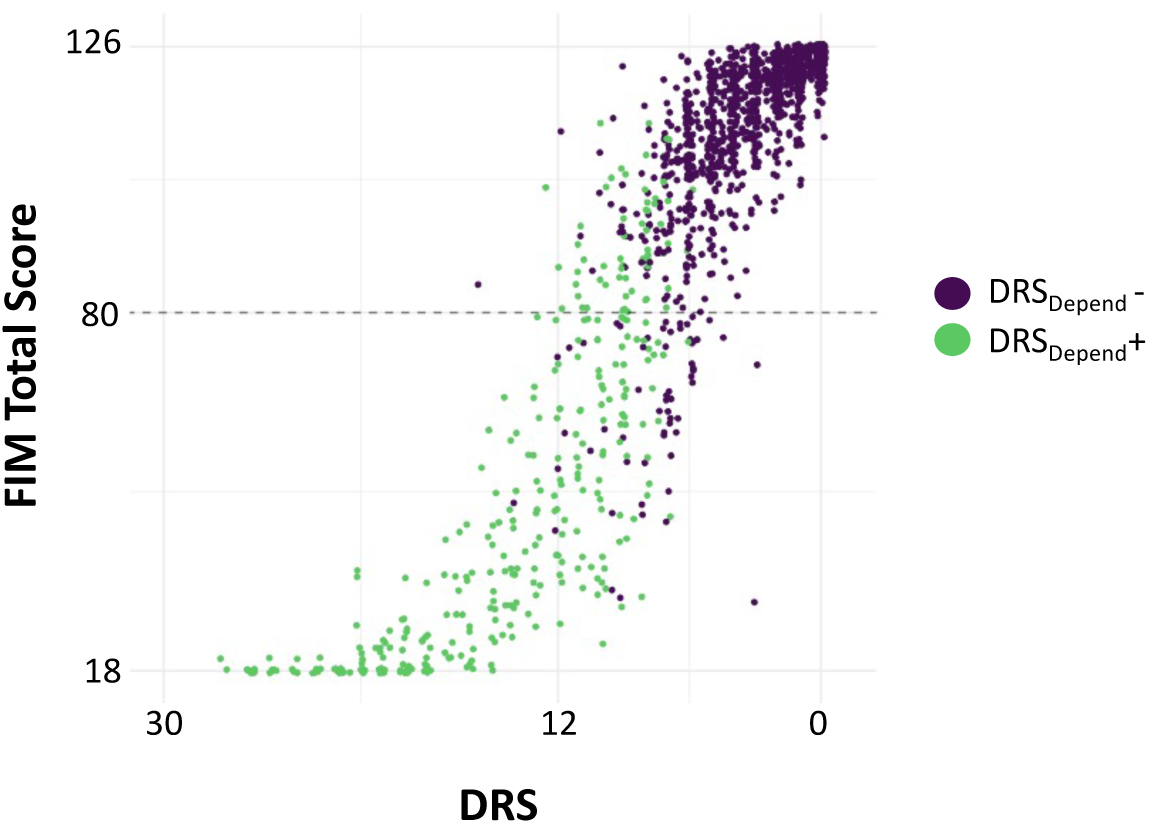
DRS_*Depend*_ Classifies Patients with FIM Scores < 80. Disability Rating Scale (DRS) scores are plotted against Functional Independence Measure (FIM) total scores. Points are randomly jittered to avoid overlap. Green color indicates the participant met criteria for DRS_Depend_, a novel DRS-based measure that seeks to identify participants dependent on others to meet basic needs, and whose impairment is at least partly due to cognitive impairment. The dashed line signifies a FIM total score of 80.

## Discussion

In this study, we examined the concordance between FIM, GOSE, and DRS scores acquired 1-year post injury in TBI survivors diagnosed with DoC on arrival to inpatient rehabilitation. We quantified the performance of literature-derived and data-driven GOSE and DRS cut-points at identifying participants who meet criteria for complete dependence on the FIM. Two common GOSE cut-points for ‘unfavorable’ outcome (i.e., ≤3 and ≤4, Lower and Upper Severe Disability, respectively) overestimated functional impairment, classifying as “dependent” 46% - 61% of participants who did not meet FIM-dependency criteria. Conversely, a DRS cut-point of Severe Disability failed to correctly identify 40% of participants meeting FIM-dependency criteria. Finally, we defined and evaluated the DRS_*Depend*_, a composite, binary rating of functional dependency due, at least in part, to cognitive impairment. We found that the DRS_*Depend*_ demonstrated better FIM-dependency classification performance than either the GOSE or DRS cut-points.

Our findings highlight the wide range of functional impairment within dichotomized GOSE and DRS categories and demonstrate fundamental differences in the calibration of these outcome scales. Most participants in this study achieved a 1-year GOSE score of 3 (Lower-Severe Disability), just one category above “Vegetative State” and typically considered an ‘unfavorable’ outcome^2, 30-35^. GOSE ≤3 was a highly sensitive but non-specific marker for FIM-dependency. Nearly all participants with FIM-dependency also had GOSE scores ≤3, but almost half of participants with GOSE ≤3 did not meet FIM-dependency criteria – with total FIM scores spanned nearly the full range of the FIM scale.

In contrast to the GOSE, the distribution of DRS scores skewed towards the milder end of the scale, suggesting a greater potential for determining meaningful functional differences between patients with DoC after TBI. However, like the GOSE ≤3 category, we found that common DRS total score groupings reported in prior studies (e.g., Moderate and Moderately-Severe)^2, 8, 37^, also spanned a wide range of FIM total scores. Although there is less precedent for dichotomizing the DRS, we found that a cut-point of at least Severe Disability (DRS ≥12) was a specific, but insensitive marker of FIM-dependency. Indeed, 40% of participants with FIM-dependency had DRS scores of Moderately-Severe Disability or better.

Our data-driven analysis identified the optimal GOSE and DRS cut-points for identifying participants meeting criteria for FIM-dependency. The optimal GOSE cut-point was ≤3, a threshold commonly used in TBI studies^2, 30-32^. Because even this optimal cut-point has a PPV only around 50% for FIM-dependency, dichotomizing outcomes using the GOSE should be done cautiously in studies enrolling severely brain injured patients. We found that the data-derived optimal DRS cut-point of ≥10 did not have an overall better discriminative performance (as measured by the AUROC) than GOSE ≤ 3. However, the higher specificity and PPV of the DRS cutpoint relative to the GOSE results in fewer participants falsely characterized as having complete dependency.

Finally, we derived and evaluated the DRS_Depend_, a novel metric that identifies participants who are dependent on others to meet basic needs, and whose impairment is at least partly due to cognitive impairment. The DRS_Depend_ outperformed data-derived optimal GOSE and DRS cutpoints at identifying participants with FIM-dependency. External validation, as well as caregiver and patient perspectives on whether the DRS_Depend_ accurately discriminates between acceptable and unacceptable outcomes^9^, requires further investigation.

### Limitations

Our findings should be interpreted in the context of several limitations. First, there is no internationally-accepted gold standard for quantifying disability and defining functional dependency. We chose the FIM as our reference standard because it is associated with daily hours of required functional assistance (burden of care)^11-14, 16^, has an established cut-point (total score < 80) for defining complete dependency^15, 16^, and was previously used in this population to define dependency^1^. Nonetheless, how this FIM threshold compares to the ground truth level of impairment is unknown.

Our study included only participants enrolled in the TBIMS with DoC on admission to inpatient rehabilitation. Restricting the study to participants with DoC may have resulted in an excess of 1-year GOSE>3 scores relative to all patients with moderate or severe TBI. Analyzing such a skewed GOSE distribution may have yielded systematically lower estimates of cross-scale concordance and classification performance. In addition, whether our results generalize to individuals with DoC who do not receive inpatient rehabilitation requires further investigation.

## Conclusions

The GOSE, DRS, and FIM have markedly different score distributions in patients recovering from post-traumatic DoC at 1-year post-injury. GOSE and DRS literature-derived cut-points have either low sensitivity or low specificity for identifying participants with FIM-dependency. A novel and simple DRS-based metric of dependency, the DRS_Depend_, identifies patients meeting criteria for FIM-dependency better than GOSE or DRS total-score cut-points.

## Data Availability

All data produced in the present study are available upon reasonable request to the TBIMS study leadership.

## Acknowledgements

The contents of this manuscript were developed under a grant from the National Institute on Disability, Independent Living, and Rehabilitation Research (NIDILRR grant numbers 90DPCP0008-01-00, 90DP0039, 90DPTB0002 (Spaulding Rehabilitation Hospital and Indiana University). NIDILRR is a Center within the Administration for Community Living (ACL), Department of Health and Human Services (HHS). The contents of this manuscript do not necessarily represent the policy of NIDILRR, ACL, HHS, and you should not assume endorsement by the Federal Government.

## Notes

### Competing Interest Statement

Dr. Zafonte reported receiving royalties from Springer/Demos for the text Brain Injury Medicine as well as serving on the scientific advisory boards of Myomo, and One Care.ai.
Dr. Giacino reported receiving a grant from the National Football League outside the submitted work. All other authors report no relevant disclosures.

### Funding Statement

SBS receives funding from the American Academy of Neurology (Clinical Research Training Scholarship).
RGK receives funding from: NIH National Institute of Neurological Disorders and Stroke (LRP).
FH receives funding from National Institute on Disability, Independent Living, and Rehabilitation Research (grants 90DPTB0002, 90DPHF0006, 90DPTB0017, and 90RTEM0008); National Institutes of Health (UG3NS117844 and 1R01NS118009), PCORI UWSC9923/PCS-1604-35115; Department of Defense (W81XWH-18-1-0796); University of California- San Francisco; University of Michigan (SUBK10416CSPR-002).
SI receives funding from the Neurocritical Care Society (Research Fellowship Award).
BLE receives funding from the NIH National Institute of Neurological Disorders and Stroke (R21NS109627, RF1NS115268), NIH Directors Office (DP2HD101400), James S. McDonnell Foundation and Tiny Blue Dot Foundation.
YGB receives funding from: NIH National Institute of Neurological Disorders and Stroke (U01 NS1365885, U01-NS086090), National Institute on Disability, Independent Living and Rehabilitation Research (NIDILRR), Administration for Community Living (90DPCP0008-01-00, 90DP0039), James S. McDonnell Foundation, and Tiny Blue Dot Foundation.

### Author Declarations

Institutional Review Board of Mass General Brigham gave ethical approval for this work.

## References

1. Kowalski RG, Hammond FM, Weintraub AH, et al. Recovery of Consciousness and Functional Outcome in Moderate and Severe Traumatic Brain Injury. JAMA Neurol 2021;78:548–557.

2. McCrea MA, Giacino JT, Barber J, et al. Functional Outcomes Over the First Year After Moderate to Severe Traumatic Brain Injury in the Prospective, Longitudinal TRACK-TBI Study. JAMA Neurol 2021;78:982–992.

3. Hammond FM, Giacino JT, Nakase Richardson R, et al. Disorders of Consciousness due to Traumatic Brain Injury: Functional Status Ten Years Post-Injury. J Neurotrauma 2019;36:1136–1146.

4. Whyte J, Giacino JT, Heinemann AW, et al. Brain Injury Functional Outcome Measure (BI-FOM): A Single Instrument Capturing the Range of Recovery in Moderate-Severe Traumatic Brain Injury. Arch Phys Med Rehabil 2021;102:87–96.

5. Nakase-Richardson R, Whyte J, Giacino JT, et al. Longitudinal outcome of patients with disordered consciousness in the NIDRR TBI Model Systems Programs. J Neurotrauma 2012;29:59–65.

6. Wilson JT, Pettigrew LE, Teasdale GM. Structured interviews for the Glasgow Outcome Scale and the extended Glasgow Outcome Scale: guidelines for their use. J Neurotrauma 1998;15:573–585.

7. Horton L, Rhodes J, Wilson L. Randomized Controlled Trials in Adult Traumatic Brain Injury: A Systematic Review on the Use and Reporting of Clinical Outcome Assessments. J Neurotrauma 2018;35:2005–2014.

8. Rappaport M, Hall KM, Hopkins K, Belleza T, Cope DN. Disability rating scale for severe head trauma: coma to community. Arch Phys Med Rehabil 1982;63:118–123.

9. Rutz Voumard R, Kiker WA, Dugger KM, et al. Adapting to a New Normal After Severe Acute Brain Injury: An Observational Cohort Using a Sequential Explanatory Design. Crit Care Med 2021;49:1322–1332.

10. Ottenbacher KJ, Hsu Y, Granger CV, Fiedler RC. The reliability of the functional independence measure: a quantitative review. Arch Phys Med Rehabil 1996;77:1226–1232.

11. Corrigan JD, Smith-Knapp K, Granger CV. Validity of the functional independence measure for persons with traumatic brain injury. Arch Phys Med Rehabil 1997;78:828–834.

12. Granger CV, Divan N, Fiedler RC. Functional assessment scales. A study of persons after traumatic brain injury. Am J Phys Med Rehabil 1995;74:107–113.

13. Dodds TA, Martin DP, Stolov WC, Deyo RA. A validation of the functional independence measurement and its performance among rehabilitation inpatients. Arch Phys Med Rehabil 1993;74:531–536.

14. Kidd D, Stewart G, Baldry J, et al. The Functional Independence Measure: a comparative validity and reliability study. Disabil Rehabil 1995;17:10–14.

15. Reistetter TA, Graham JE, Deutsch A, Granger CV, Markello S, Ottenbacher KJ. Utility of functional status for classifying community versus institutional discharges after inpatient rehabilitation for stroke. Arch Phys Med Rehabil 2010;91:345–350.

16. Uniform Data System for Medical Rehabilitation. The FIM Instrument: its background, structure, and usefulness Published 2012 accessed October 1, 2021. http://docshare02.docshare.tips/files/23187/231876238.pdf

17. Black TM, Soltis T, Bartlett C. Using the Functional Independence Measure instrument to predict stroke rehabilitation outcomes. Rehabil Nurs 1999;24:109-114, 121.

18. Chumney D, Nollinger K, Shesko K, Skop K, Spencer M, Newton RA. Ability of Functional Independence Measure to accurately predict functional outcome of stroke-specific population: systematic review. J Rehabil Res Dev 2010;47:17–29.

19. Bagiella E, Novack TA, Ansel B, et al. Measuring outcome in traumatic brain injury treatment trials: recommendations from the traumatic brain injury clinical trials network. J Head Trauma Rehabil 2010;25:375–382.

20. McMillan T, Wilson L, Ponsford J, Levin H, Teasdale G, Bond M. The Glasgow Outcome Scale - 40 years of application and refinement. Nat Rev Neurol 2016;12:477–485.

21. Zuckerman DA, Giacino JT, Bodien YG. Traumatic Brain Injury: What Is a Favorable Outcome? J Neurotrauma 2021.

22. Gouvier WD, Blanton PD, LaPorte KK, Nepomuceno C. Reliability and validity of the Disability Rating Scale and the Levels of Cognitive Functioning Scale in monitoring recovery from severe head injury. Arch Phys Med Rehabil 1987;68:94–97.

23. Traumatic Brain Injury Model Systems National Data and Statistical Center. Welcome to the NDSC accessed September 4, 2021. https://www.tbindsc.org/

24. Corrigan JD, Cuthbert JP, Whiteneck GG, et al. Representativeness of the Traumatic Brain Injury Model Systems National Database. J Head Trauma Rehabil 2012;27:391–403.

25. Tso S, Saha A, Cusimano MD. The Traumatic Brain Injury Model Systems National Database: A Review of Published Research. Neurotrauma Rep 2021;2:149–164.

26. Traumatic Brain Injury Model Systems National Data and Statistical Center. Standardized operating procedure 101a: identification of subjects for the TBI Model Systems National Database accessed October 1, 2021. https://www.tbindsc.org/StaticFiles/SOP/101a%20-%20Identification%20of%20Subjects.pdf

27. Pettigrew LE, Wilson JT, Teasdale GM. Assessing disability after head injury: improved use of the Glasgow Outcome Scale. J Neurosurg 1998;89:939–943.

28. Maas AI, Harrison-Felix CL, Menon D, et al. Common data elements for traumatic brain injury: recommendations from the interagency working group on demographics and clinical assessment. Arch Phys Med Rehabil 2010;91:1641–1649.

29. Weir J, Steyerberg EW, Butcher I, et al. Does the extended Glasgow Outcome Scale add value to the conventional Glasgow Outcome Scale? J Neurotrauma 2012;29:53–58.

30. Wilkins TE, Beers SR, Borrasso AJ, et al. Favorable Functional Recovery in Severe Traumatic Brain Injury Survivors beyond Six Months. J Neurotrauma 2019;36:3158–3163.

31. Hutchinson PJ, Kolias AG, Timofeev IS, et al. Trial of Decompressive Craniectomy for Traumatic Intracranial Hypertension. N Engl J Med 2016;375:1119–1130.

32. Vahedi K, Hofmeijer J, Juettler E, et al. Early decompressive surgery in malignant infarction of the middle cerebral artery: a pooled analysis of three randomised controlled trials. Lancet Neurol 2007;6:215–222.

33. Yuh EL, Jain S, Sun X, et al. Pathological Computed Tomography Features Associated With Adverse Outcomes After Mild Traumatic Brain Injury: A TRACK-TBI Study With External Validation in CENTER-TBI. JAMA Neurol 2021;78:1137–1148.

34. Okonkwo DO, Shutter LA, Moore C, et al. Brain Oxygen Optimization in Severe Traumatic Brain Injury Phase-II: A Phase II Randomized Trial. Crit Care Med 2017;45:1907–1914.

35. Cooper DJ, Nichol AD, Bailey M, et al. Effect of Early Sustained Prophylactic Hypothermia on Neurologic Outcomes Among Patients With Severe Traumatic Brain Injury: The POLAR Randomized Clinical Trial. JAMA 2018;320:2211–2220.

36. Teasdale G, Jennett B. Assessment of coma and impaired consciousness. A practical scale. Lancet 1974;2:81–84.

37. Giacino JT, Whyte J, Bagiella E, et al. Placebo-controlled trial of amantadine for severe traumatic brain injury. N Engl J Med 2012;366:819–826.

38. Wilson L, Boase K, Nelson LD, et al. A Manual for the Glasgow Outcome Scale-Extended Interview. J Neurotrauma 2021;38:2435–2446.

39. Christian Thiele GH. cutpointr: Improved Estimate and Validation of Optimal Cutpoints in R. Journal of Statistical Software 2021;98.

