## Supplementary Material for "Comparison of common outcome measures for assessing independence in patients diagnosed with disorders of consciousness: A Traumatic Brain Injury Model Systems Study"

### SUPPLEMETARY MATERIAL

Samuel B. Snider MD[1], Robert G. Kowalski, MBBCh, MS[2], Flora Hammond MD[3], Saef Izzy MD[1], Shirley L. Shih MD[4], Craig Rovito MD[4], Brian L. Edlow MD[5], Ross D. Zafonte DO[4], Joseph T. Giacino PhD[4], Yelena G. Bodien PhD[4,5]

**Affiliations**

1 Division of Neurocritical Care, Department of Neurology, Brigham and Women's Hospital, Boston MA, USA

2 Departments of Neurosurgery and Neurology, University of Colorado School of Medicine, Aurora CO

3 Department of Physical Medicine and Rehabilitation, Indiana University School of Medicine, Indianapolis, IN, USA

4 Department of Physical Medicine and Rehabilitation, Spaulding Rehabilitation Hospital and Harvard Medical School, Charlestown, MA

5 Center for Neurotechnology and Neurorecovery, Department of Neurology, Massachusetts General Hospital and Harvard Medical School, Boston, MA

| Supplementary Table 1: FIM-dependency classification performance measures | | |
|---|---|---|
| **Measure** | **Definition** | **Explanation in context of GOSE ≤ 3 cutpoint** |
| Sensitivity | $\frac{TP}{TP + FN}$ | % of patients with FIM < 80 who are GOSE ≤ 3 |
| Specificity | $\frac{TN}{TN + FP}$ | % of patients with FIM ≥ 80 who are GOSE > 3 |
| Positive Predictive Value | $\frac{TP}{TP + FP}$ | % of patients with GOSE ≤ 3 who have FIM < 80 |
| Negative Predictive Value | $\frac{TN}{TN + FN}$ | % of patients with GOSE > 3 who have FIM ≥ 80 |

**Terms/Abbreviations:**

TP = True Positive: GOSE ≤ 3 & FIM < 80
FP = False Positive: GOSE ≤ 3 & FIM ≥ 80
TN = True Negative: GOSE > 3 & FIM ≥ 80
FN = False Negative: GOSE > 3 & FIM < 80
